# Distance from home to motor vehicle crash location: Implications for license restrictions among medically-at-risk older drivers

**DOI:** 10.1101/2021.08.20.21262345

**Authors:** Nina R. Joyce, Marzan A. Khan, Andrew R. Zullo, Melissa R. Pfeiffer, Kristina B. Metzger, Seth A. Margolis, Brian R. Ott, Allison E. Curry

**Affiliations:** Department of Epidemiology, Brown University School of Public Health; Center for Gerontology and Health Care Research, Brown University School of Public Health; Department of Health Services Policy and Practice, Brown University School of Public Health; Center of Innovation in Long-term Services and Supports, Providence Veterans Affairs Medical Center, Providence, Rhode Island; Center for Injury Research and Prevention, Children’s Hospital of Philadelphia; Rhode Island Hospital, Providence, Rhode Island; Department of Psychiatry & Human Behavior, Brown University; Department of Neurology, Brown University; Division of Emergency Medicine, Perelman School of Medicine, University of Pennsylvania

**Keywords:** chronic disease, traffic accidents, Medicare, motor vehicles, elderly, driving, automobile, living, independent, traffic collisions, transportation, Activities of Daily Living

## Abstract

**Background/Objectives:** Thirty states allow licensing agencies to restrict the distance from home that “medically-at-risk” drivers are permitted to drive. However, there is little information on where older drivers crash relative to their home or how distance to crash varies by medical condition, and thus, what impact distance limits may have on motor vehicle crash rates for “medically-at-risk” drivers.

**Design:** Observational study of crash-involved drivers.

**Setting:** Medicare fee-for-service claims linked to geocoded crash locations and residential addresses from police crash reports in the state of New Jersey from 2007 through 2017.

**Participants:** New Jersey Medicare fee-for-service beneficiaries aged 68 years and older involved in police-reported crashes.

**Measurements:** The outcome was Euclidian distance from home to crash location. Covariates included driving-relevant medical conditions from Medicare claims, crash characteristics from police reports, and demographics from both sources.

**Results:** There were 197,122 crash-involved older drivers for whom approximately 70% of crashes occurred within 5 miles and 95% within 25 miles of the driver’s residence. The mean distance to crash was 6.0 miles. Although distance from home to the crash was generally lower among drivers with (versus without) each of the medical conditions studied, the differences were small (maximum mean difference of 2.1 miles). The largest difference in distance was by licensure status, where unlicensed/suspended drivers crashed significantly farther from home than validly licensed drivers (8.8 miles, 95% Confidence Interval [CI]: 8.4-9.1 vs 5.9 miles, 95% CI: 5.9 – 6.0).

**Conclusions:** Findings suggest that the majority of older adults who crash do so within a few miles from home and that the distance to crash does not differ substantially by the presence of a driving-relevant medical condition. Thus, distance restrictions may not reduce crash rates among older adults and the tradeoff between safety and mobility warrants consideration.

**KEYPOINTS:**

- 30 states permit restriction of driving distance for medically-at-risk drivers
- The vast majority of crashes involving older drivers occur ≤ 25 miles of home
- Distance to crash does not differ markedly across driving-relevant medical conditions

**Why does this matter?:**

- Distance restrictions are likely to impact mobility and autonomy but not crash rates

## INTRODUCTION

By 2050, the US population 65 years and older is expected to almost double to 84 million people, the vast majority of whom are expected to be licensed to drive.^1^ Consequently, there is growing interest in interventions that reduce the risk of crash in older driver populations without negatively impacting the independence that driving provides.^2^ Driver’s license restrictions that limit the distance from home that an individual is permitted to drive may be one way to achieve this balance by allowing older drivers to access nearby essential destinations, while limiting their access to unfamiliar terrain where the risk of a crash may be greater.^3-6^ These “distance-from-home” restrictions are currently permitted in thirty states for “medically-at-risk” drivers, though the details of the policies vary.^7^ For instance, in Maine, drivers with poor visual acuity may be restricted to driving within a 25-mile radius of their home, while drivers with dementia can request an in-person neighborhood driving exam to determine the appropriate radius. In Virginia, the radius is determined by the distance to the locations the driver frequents (e.g., the doctor, the grocery store).^5^ Although these restrictions are not limited to older adults, the physical and mental conditions that define a medically-at-risk driver, such as vision loss, dementia and stroke, are more common in older adults.

“Distance-from-home” restrictions can only be effective in reducing the risk of a crash if crashes among medically-at-risk drivers are more prevalent farther from home. If, instead, crashes are more prevalent closer to home in this population, then geographic restrictions may serve to unnecessarily limit travel without substantially reducing crash risk. Currently, there is little information on the location of crashes relative to a driver’s home address, particularly for older drivers with clinical conditions that increase the risk of a crash. Available data on crash distance from home is limited to (1) insurance company surveys with no additional information about the driver or their relevant medical conditions; ^8^ (2) information from different countries where the driving environment and policy landscape may not translate easily to the US ^9,10^ and (3) only crude measures of distance, such as distance from the centroid of the of the driver’s home address census tract to the crash location.^11^

Therefore, we sought to characterize the distribution of the distance from a driver’s home address to the location of a crash and identify driver characteristics, including clinical conditions relevant for driving, associated with distance to the crash using data on all police-reported crashes among Medicare fee-for-service beneficiaries in the state of New Jersey from 2007 through 2017.

## METHODS

### Data Sources and Linkage

The data for this study came from the New Jersey Safety and Health Outcomes (NJ-SHO) data warehouse (2007-2017), the Medicare Master Beneficiary Summary File (MBSF) (2004-2017), and the Medicare Chronic Conditions Data Warehouse (CCW) (2004-2017). We developed the NJ-SHO warehouse as an extensive repository that integrates data from multiple New Jersey statewide administrative sources, including driver licensing and police-reported crash data; full details of our development of the NJ-SHO warehouse are available in prior work.^12^ Driver residence and crash location were both obtained from the police crash report.

The MBSF contains Medicare beneficiary enrollment and demographic information, while the CCW contains information on chronic and potentially disabling conditions constructed from Medicare fee-for-service insurance claims. Medicare data were linked to the NJ-SHO warehouse by a third party (General Dynamics Information Technology) using a strict matching algorithm that included last name, birthdate, sex and zip code.

### Study Population

We identified all drivers involved in police-reported crashes (i.e., crashes that led to injury or death of a person or greater than $500 in property damage) from the NJ-SHO data warehouse (Figure 1).^13^ The primary unit of analysis was the crash-involved older driver; thus, a single individual may be included in the study population more than once if they were a driver involved in multiple crashes over the study period.

**Figure 1:**
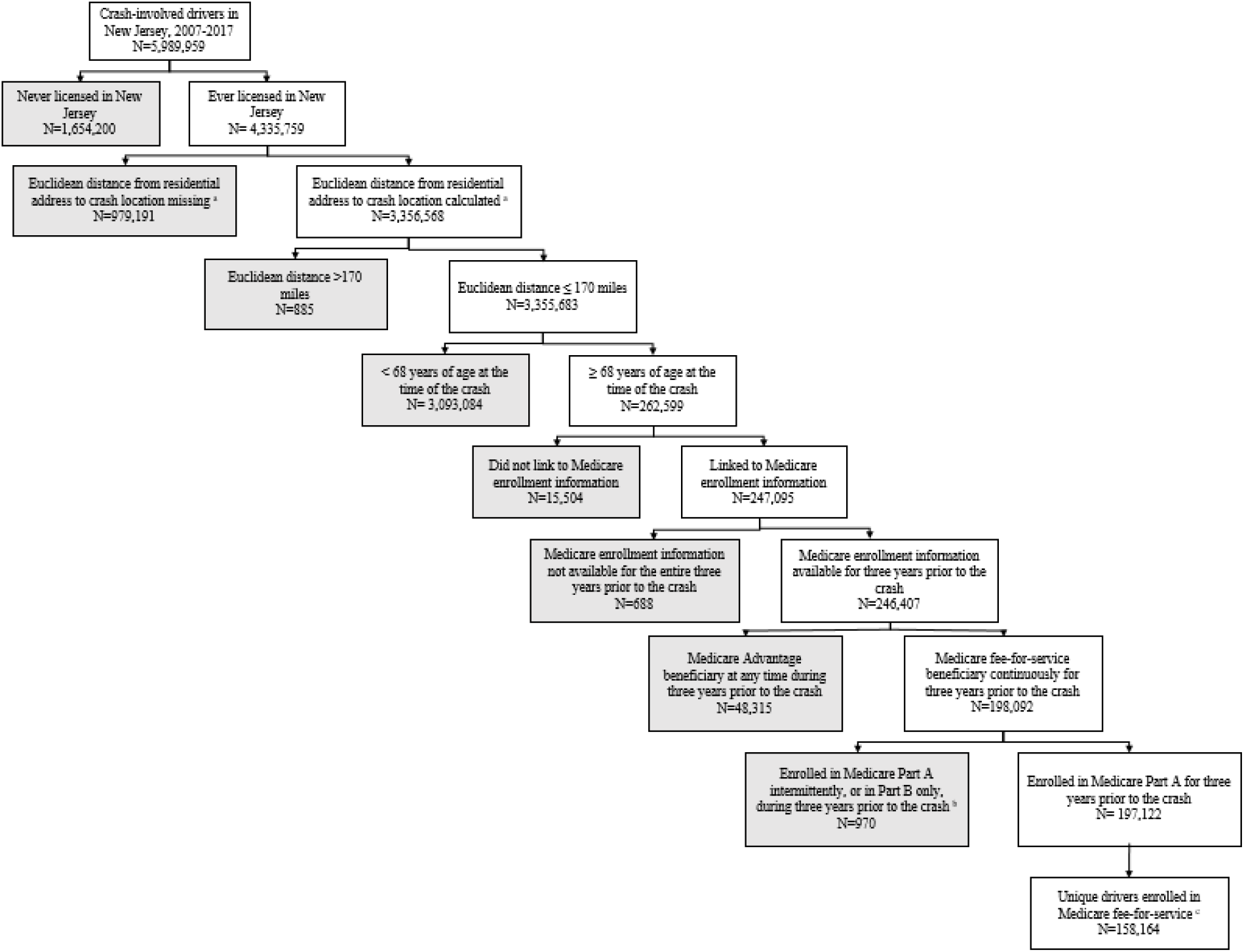
Flow chart illustrating selection of the final study sample, with the grey boxes depicting exclusions conducted. ^a^ Euclidean distance from residential address as recorded on the crash report. ^b^we excluded crash-involved drivers for whom the population density of their home address census tract was unavailable due to small-cell-size publication restriction required by the Centers for Medicare and Medicaid (CMS). ^c^A single individual may be included in the study population more than once if they were a driver involved in multiple crashes over the study period.

We excluded crash-involved drivers who were never licensed in New Jersey or did not have a full license (as it is not clear that a distance restriction would apply to them). Crash-involved drivers who had their residential address or crash location geocoded and for whom Euclidean distance (i.e., straight line distance) between their residential address and crash location were available, were included in the study sample. Further, we excluded crash-involved drivers with a Euclidean distance of greater than 170 miles, which is the maximum Euclidean distance within the state.^14-16^

We excluded all crash-involved drivers less than 68 years as well as those who could not be linked to the MBSF, were enrolled in Medicare Advantage at any time up to three years before the crash, were enrolled in Medicare Part A intermittently during the three years prior to the crash or were beneficiaries of Medicare Part B only. These restrictions were necessary to obtain complete data on clinical conditions from the CCW, which have a reference period of up to 3 years (consequently, the minimum age for inclusion into the study was 68 years). In addition, we excluded fewer than 10 crash involved drivers for whom the population density of their home census tract was unavailable due to small-cell-size publication restrictions required by the Centers for Medicare and Medicaid (CMS).

### Outcome

The outcome was the Euclidean distance (henceforth referred to as “distance”) from the residential address of a crash-involved driver to their crash location. Residential addresses were geocoded using ArcGIS V.10.5 (Esri, Redlands, California, USA). Records that consisted of at least one element of the address (i.e., street, city, state, zip code) and that were associated with the state of NJ or an unknown state were prepared for geocoding. The default geocoding options such as spelling sensitivity (80), minimum candidate score (75) and minimum match score (85) were utilized during the geocoding process and the results were compared against Google Maps and included coordinate values (latitude and longitude) and census tract. We conducted a hand review of 500 randomly sampled records to assess the success of the geocoding of home addresses; results estimated the true match rate to be 99.7%. Crash locations were geocoded by the NJ Department of Transportation.^12^

### Driver and Crash Measures

Driver age at the time of the crash, licensing status, census tract population density, crash-related fatality and crash characteristics were determined from the crash data contained in the NJ-SHO warehouse. The 2013-2017 American Community Survey 5-year population estimates and the geographic area in square miles from the 2010 Census Gazetteer Files were used to derive quintiles of population density (population per square miles) for the NJ census tracts.^17,18^

Driver sex and race/ethnicity were ascertained from the MBSF while clinical conditions were determined from the CCW. The definition of “medically-at-risk” can vary across states, and thus we included conditions in the CCW that were suggested by the “Clinician’s Guide to Assessing and Counseling Older Drivers” developed by The American Geriatrics Society and the National Highway Traffic Safety Administration as potentially driving-relevant conditions. ^19^ Each condition was analyzed as a dichotomous variable distinguishing those who were ever, as compared to never, diagnosed prior to the date of the crash based on the earliest date of their diagnosis in the CCW. Information on condition severity was not available. See **Supplementary Table S1** for a complete list of the clinical conditions.

### Statistical Analysis

We plotted the cumulative proportion of crashes by distance (rounded up to the nearest integer) from residential address to the crash location. We calculated the mean distance from residence to the crash location and the associated 95% confidence interval (CI) by driver characteristics and clinical conditions. We used robust standard errors to account for multiple crashes per driver. In addition, we calculated the difference in mean distance to crash for crash-involved drivers with, compared to without, the clinical conditions using a two-sample t-test.

### Software

All analyses were conducted using SAS software, Version 9.4 (SAS Institute Inc., Cary, NC). This analysis was reviewed and approved by the Brown University Institutional Review Board.

## RESULTS

The analytic sample included 197,122 crash-involved drivers (**Figure 1**). This sample included 158,164 unique drivers who were involved in one or more crashes during the study period (Table 1). Crash-involved drivers had a mean age of 76.1 years (Standard Deviation = 6.3), were predominantly Non-Hispanic White (83.3%) and slightly more likely to be male (53.0%) (**Table 1**). Over one-third of crash-involved drivers were 68-72 years of age (36.7% and, a minority were 93-103 years old (0.7%). A majority of drivers were involved in only one crash from 2007 through 2017 (81.2%) and held a valid license at the time of the crash (97.5%). The second quintile of population density (1,299 - 2,919 population per square mile) constituted the largest proportion of crash-involved drivers (27.2%). Fatalities among crash-involved drivers were uncommon (0.2%). Among the clinical conditions present in the sample, history of cataracts (63.9%), ischemic heart disease (55.0%), rheumatoid arthritis (51.6%), diabetes (40.7%), glaucoma (27.0%) and heart failure (26.3%) were the most prevalent (**Supplementary table S1**). Less common conditions included visual impairment (0.2%), Attention-Deficit/Hyperactivity Disorder (ADHD) and other conduct disorders (0.3%), and personality disorders (0.5%). Alzheimer’s disease and related disorders and senile dementia was present in 7.8% of crash-involved drivers.

**Table 1:** Characteristics of crash-involved drivers in New Jersey, ages 68 years and older, 2007-2017.

Characteristics
|  |  |
| --- | --- |
| **Crash-involved driver level <sup>a,b</sup>** | **n=197,122** |
| Age, mean (SD), years | 76.1 (6.3) |
| Age group, N (%) |  |
| 68-72 years | 72,407 (36.7) |
| 73-77 years | 50,976 (25.9) |
| 78-82 years | 38,144 (19.4) |
| 83-87 years | 25,011 (12.7) |
| 88-92 years | 9,124 (4.6) |
| 93-103 years | 1,460 (0.7) |
| License status, N (%) |  |
| Valid driver's license | 192,107 (97.5) |
| Unlicensed or suspended driver's license | 5,015 (2.5) |
| Driver fatality in crash, N (%) <sup>c</sup> |  |
| Driver died | 453 (0.2) |
| Quintile of population density, population per square mile, N (%) |  |
| 1st: < 1,299 | 44,842 (22.7) |
| 2nd: 1,299 - 2,919 | 53,651 (27.2) |
| 3rd: 2,920 - 5,237 | 44,068 (22.4) |
| 4th: 5,238 - 12,711 | 35,954 (18.2) |
| 5th: ≥ 12,712 | 18,607 (9.4) |
| **Driver level <sup>a</sup>** | **n=158,164** |
| Sex, N (%) |  |
| Male | 83,858 (53.0) |
| Female | 74,306 (47.0) |
| Race/Ethnicity, N (%) |  |
| Non-Hispanic White | 131,759 (83.3) |
| Black or (African American) | 12,763 (8.1) |
| Hispanic | 6,624 (4.2) |
| Asian/ Pacific Islander | 4,646 (2.9) |
| Other | 1,662 (1.1) |
| Unknown | 672 (0.4) |
| American Indian/ Alaskan Native | 38 (0.0) |
| Number of crashes in study period, N (%) |  |
| 1 | 128,447 (81.2) |
| 2 | 23,155 (14.6) |
| 3 | 4,783 (3.0) |
| ≥ 4 | 1,779 (1.1) |

SD=standard deviation, %=percentage
<sup>a</sup> Because a driver could be involved in more than one crash, crash-involved-driver level information refers to measures that could vary between crashes, whereas driver level information refers to measures that were time-invariant (sex, race) or was a summary over the study period (number of crashes).
<sup>b</sup> All measures reflect the status at the time of the crash.
<sup>c</sup> A driver is considered to have died in the crash if their death occurred on the same day or within 7 days of the crash. The frequency and proportion of drivers who did not die in the crash are not shown in the table.

### Overall Distance to Crash

The overall mean distance to crash among older drivers was 6.0 miles. We plotted the cumulative proportion of crashes by distance to crash using a bar graph in **Figure 2**. A steep increase in the cumulative proportion of crashes was observed within the first three miles (27.2% of the crashes occurred within 1 mile, 44.9% occurred within 2 miles and 56.4% occurred within 3 miles), which was followed by more gradual increments. Approximately 95% of the crashes occurred within 25 miles from drivers’ residences; the maximum distance was 152 miles (**Figure 2**).

**Figure 2:**
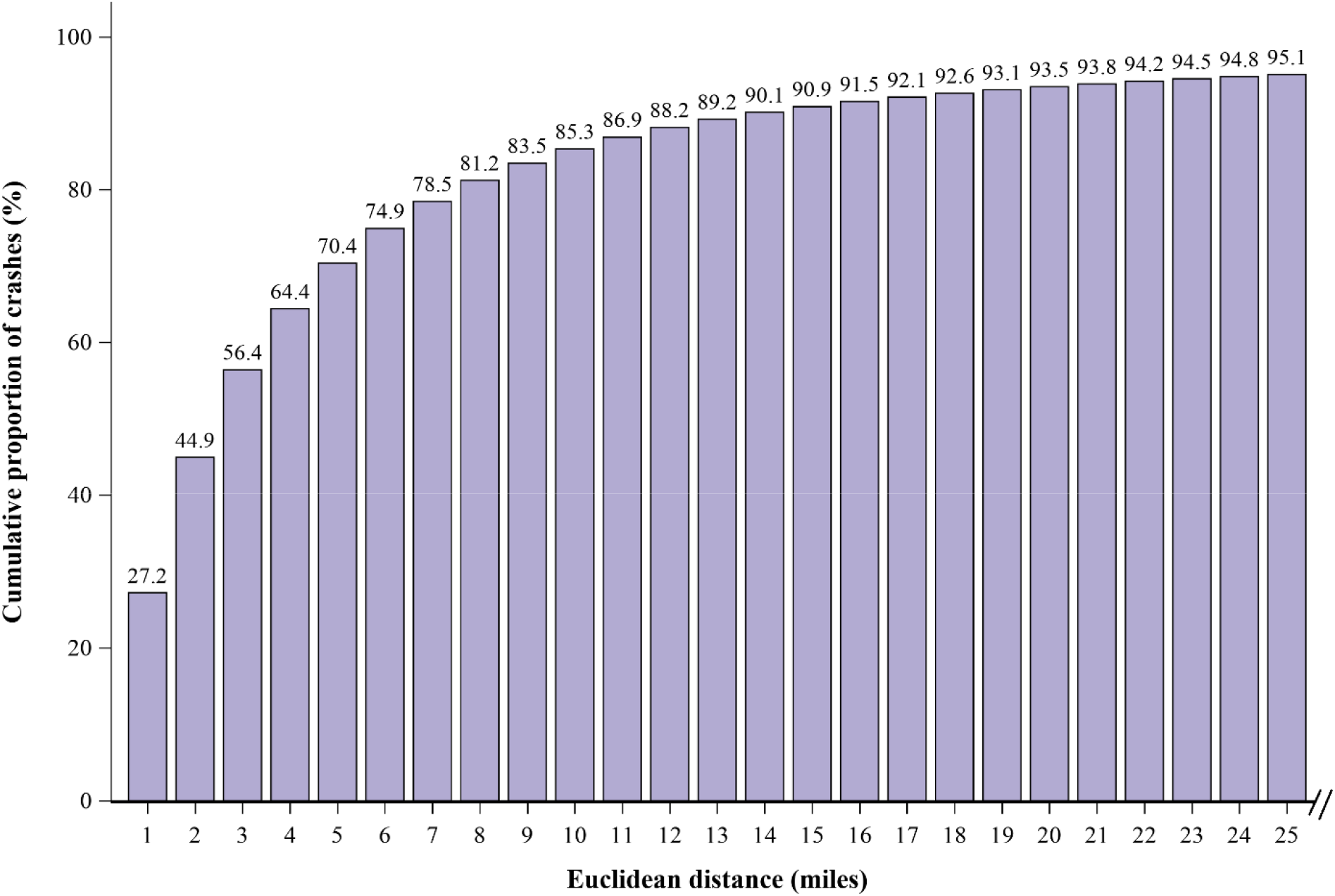
Cumulative proportion of crashes by Euclidean distance from residential address to the crash location among crash-involved drivers in New Jersey, ages 68 years and older, 2007-2017. Note: 95% of the crashes occurred within 25 miles from the driver’s residence. The maximum value of Euclidean distance in the data was 152 miles.

### Distance to Crash by Demographic Characteristics

The mean Euclidean distance to crash decreased with age (age group 68 to 72 years: mean=7.1 miles, 95% CI=7.0 – 7.2 vs age group 93 to 103 years: mean=3.3 miles, 95% CI=2.9 – 3.7). Males crashed farther from their homes on average compared to females (mean=6.8, 95% CI=6.7 – 6.9 vs. mean=5.0 miles, 95% CI=5.0 – 5.1, respectively). Crash-involved drivers who were unlicensed or had a suspended license or who died in the crash crashed farther away from their homes on average compared to their respective counterparts. Crash-involved drivers who belonged to the first quintile of population density (< 1,299 population per square mile) crashed farthest away from their homes compared to others. The largest difference in the distance to crash across all of the driver characteristics studied (including clinical condition) was by licensure status, with unlicensed/suspended drivers crashing farther from home (**Figure 3**).

**Figure 3:**
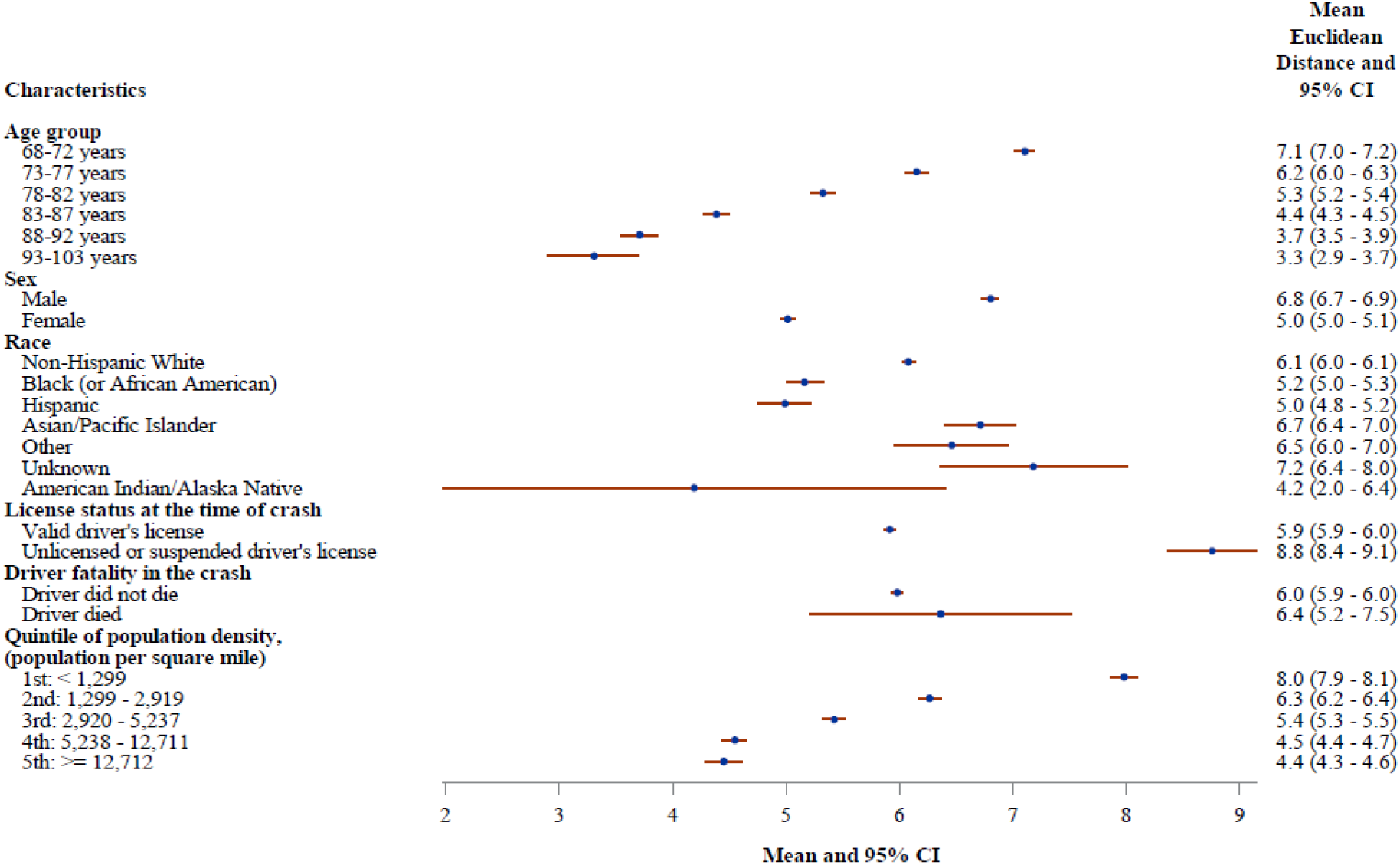
Forest plot of mean Euclidean distance from residential address to the crash location and associated 95% confidence intervals by characteristics of the crash-involved drivers in New Jersey, ages 68 years and older, 2007-2017. CI = Confidence Interval

### Distance to Crash by Clinical Conditions

The mean distance to crash varied across the clinical conditions of the drivers, with values ranging from 3.9 miles (95% CI=3.2 - 4.6) among drivers with sensory blindness and visual impairment to 6.4 miles (95% CI=5.5 - 7.3) among those with ADHD and other conduct disorders (**Figure 4**). For each condition, drivers with the condition tended to crash closer to their homes compared to those without the condition, except for those with ADHD and other conduct disorders and those with personality disorders. The greatest mean difference in distance to crash was 2.1 miles (95% CI=-2.8 – -1.4) comparing drivers diagnosed with sensory blindness and visual impairment to those who were not (**Supplementary figure S1**).

**Figure 4:**
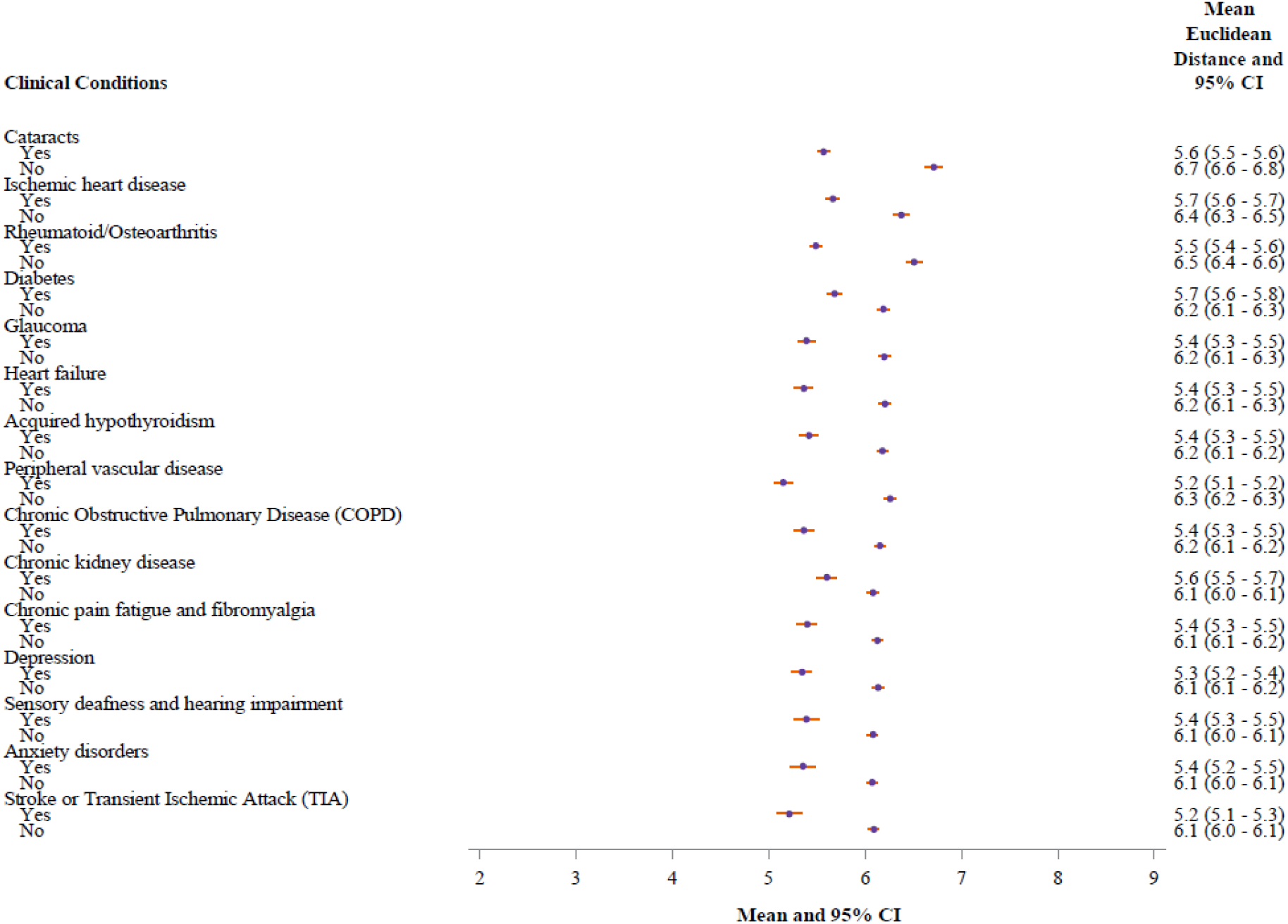
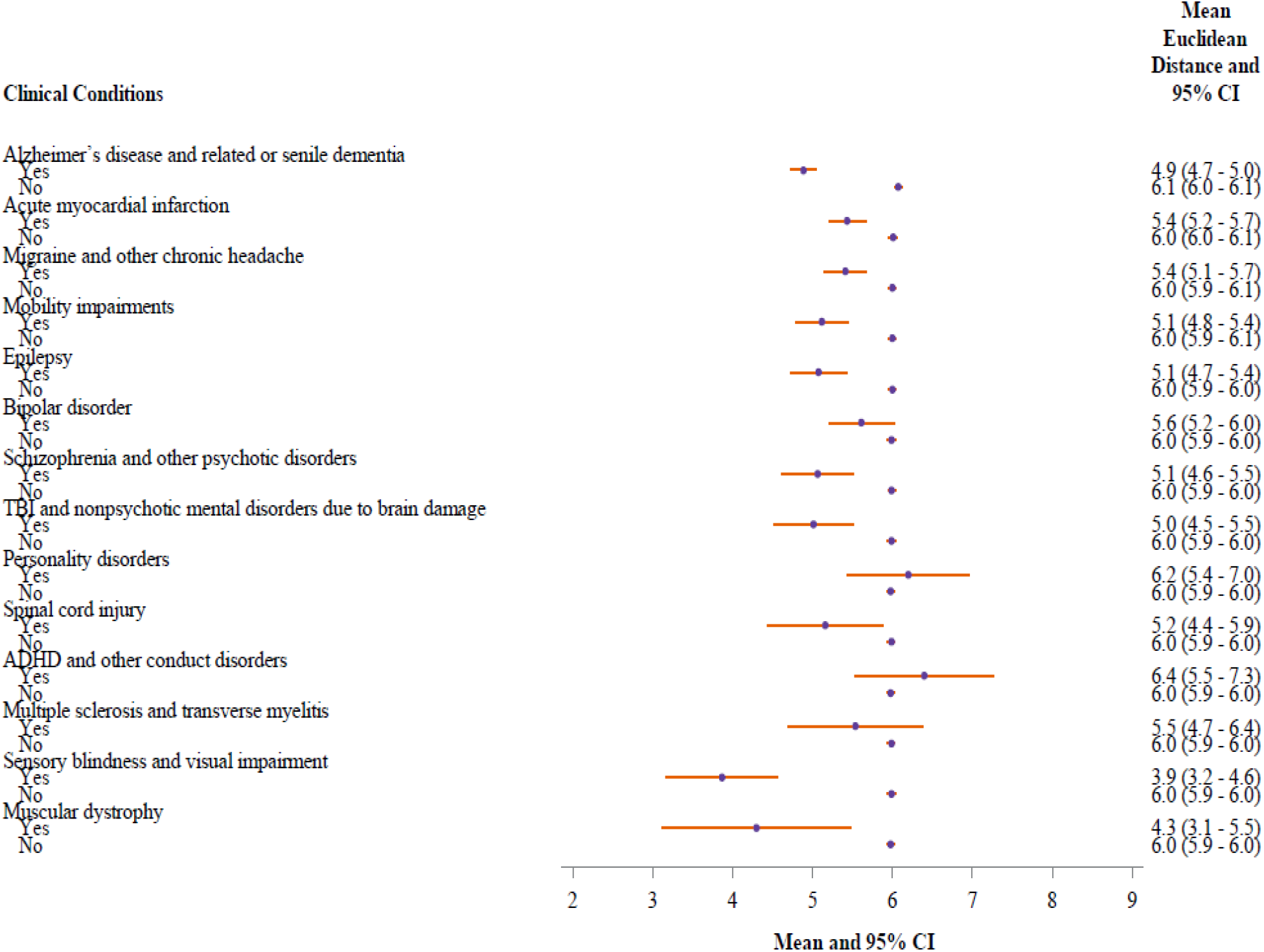
Forest plot of mean Euclidean distance from residence to crash location and associated 95% confidence intervals by comorbidities of drivers in New Jersey, ages 68 years and older, 2007-2017. CI = Confidence Interval, TBI=Traumatic Brain Injury, ADHD=Attention-Deficit/Hyperactivity Disorder

## DISCUSSION

As the number of older drivers continues to rise, the need to balance mobility and autonomy with their safety and the safety of those with whom they share the road has become increasingly important. License restrictions, such as “distance-from-home” limits, are one way that states may be able to meet these dual needs. To be effective, however, distance restrictions need to target locations where the risk of crash is highest. In this study of over 190,000 crash-involved drivers 68 years and older, we found that the overwhelming majority (95%) of crashes occurred within 25 miles of the driver’s residence. While there were small but notable patterns in the mean distance to crash location by clinical characteristics of the driver, the maximum mean distance from home for a particular group never exceeded nine miles. Thus, our findings suggest that irrespective of clinical diagnoses, older adults tend to crash close to their home and, as a result, distance restrictions will likely have little impact on crash rates among older drivers.

Although the mean difference in the distance to crash by clinical condition was quite small, there was a consistent pattern in that, apart from ADHD and personality disorders, crash-involved drivers with the condition crashed closer to home on average than those without the condition. This pattern may reflect the tendency of older drivers with perceived limitations to self-regulate by driving closer to their home, in areas where they feel more comfortable.^20-22^ For instance, a survey of older adults recruited from a memory clinic found that those diagnosed with Alzheimer’s Disease and Related Dementias (ADRD) were significantly more likely to limit both the frequency of driving as well as the conditions under which they would drive (i.e., avoiding driving at night and on unfamiliar roads).^21^ Further, in one naturalistic driving study, patients with Alzheimer’s disease displayed further restrictions of driving behavior beyond those of healthy older adults, suggesting additional regulation on the basis of cognitive status.^23^ For these drivers, restrictions placed by family members and other caregivers likely also played a role.^24^

Self-regulation has been described as a risk reduction strategy that involves a more complex interaction of factors beyond health status alone and includes age and sex, as well as driving confidence in general.^25^ To this end, our finding that age and sex were both associated with distance to crash is consistent with prior literature on self-regulation of driving among older adults. Numerous studies have shown that as adults age they self-limit their driving, both the duration and the conditions in which they drive, and that this is true even among adults without medical conditions that may put them at greater risk of a crash.^22,26^ Similarly, several studies have shown that, compared to men, women are more likely to self-limit their driving, even when they are considered safe to continue driving. ^26,27 28^

Our study had several limitations worth discussing. First, our data are from a single state, New Jersey, and thus may not reflect the distribution of the distance from home to the crash location in other states, particularly other states with a different distribution of population density. However, we also view this as a strength since New Jersey does not have a “distance-from-home” license restriction and therefore our findings are unaffected by policies that already limit where older adults can drive. Second, we limited our sample to continuously enrolled Medicare fee-for-service beneficiaries, and thus our findings may not apply to beneficiaries participating in Medicare Advantage or those with Part B coverage only (without Part A). Third, we classified older drivers as having a condition if they received the diagnosis at any point prior to the crash date. Though some conditions are chronic and persistent (e.g., stroke), others are progressive (e.g., ADRD), and some may be transient or can be treated (e.g., cataracts). Consequently, for some of the conditions studied it is possible that the driver did not have that condition at the time of the crash. Furthermore, because our data only contain information on the presence or absence of a diagnosis, we were also unable to account for differences in severity of illness. Last, we are unable to determine if older adults are more likely to crash closer to their home because that is where they spend the most time driving, or if there is something riskier about the driving conditions close to home. While distance restrictions are meant to reduce crash risk by limiting driving to familiar territory, some researchers have suggested that crash risk may be higher closer to home because this familiarity leads to overconfidence.^9^ However, without additional information on driving exposure (e.g., crashes per mile driven) it is not possible to determine whether restrictions according to distance versus number of trips or miles driven would be the most effective approach to reduce crash frequency.

## CONCLUSION

License restrictions have become an increasingly common policy tool for lowering crash rates, particularly among older drivers. Both the National Highway Traffic Safety Administration (NHTSA) and The American Automobile Association (AAA) have included restricted license as one of their recommendations towards safe mobility among older adults.^6,19^ “Distance-from-home” restrictions are meant to address the risk of crash among medically-at-risk drivers by limiting their driving to familiar surroundings and may be an important tool for balancing safety with the mobility needs of older adults.^6^ However, given that the overwhelming majority of older adult crashes occur close to home, polices that restrict the distance “medically-at-risk” drivers can drive relative to their home will have little impact on crash risk in this population. Additional work is needed to determine crash risk according to driving exposure at various distances from home as well as the implications of distance restrictions for mobility among older drivers

## Data Availability

The data are not publicly available

## ACKNOWLEDGEMENTS

The authors would like to acknowledge Heather Griffis, PhD and Vicky Tam, MA for their efforts in geocoding activities.

## Summary of conflict of interest statements

No conflicts to declare.

The authors confirm the following contribution to the paper as follows:

1. study conception and design: ARZ, NRJ, AEC, KBM
2. Analysis and interpretation: MAK, ARZ, NRJ, AEC, KBM, MRP, SM, BO
3. Draft manuscript preparation: MAK, ARZ, NRJ
4. Critical revisions: MAK, ARZ, NRJ, AEC, KBM, MRP, SM, BO

