## Supplementary Tables and Figures for "Distance from home to motor vehicle crash location: Implications for license restrictions among medically-at-risk older drivers"

**Supplementary Table S1:** Frequency and proportion of clinical conditions among crash-involved drivers in New Jersey, ages 68 years and older, 2007-2017 (n=197,122).

| **Clinical conditions** [a] | **N (%)** |
|---|---|
| Cataracts | 125,865 (63.9) |
| Ischemic heart disease | 108,452 (55.0) |
| Rheumatoid/Osteoarthritis | 101,622 (51.6) |
| Diabetes | 80,284 (40.7) |
| Glaucoma | 53,272 (27.0) |
| Heart failure | 51,814 (26.3) |
| Acquired hypothyroidism | 51,090 (25.9) |
| Peripheral vascular disease | 49,592 (25.2) |
| Chronic Obstructive Pulmonary Disease (COPD) | 42,983 (21.8) |
| Chronic kidney disease | 39,549 (20.1) |
| Chronic pain fatigue and fibromyalgia | 38,906 (19.7) |
| Depression | 37,135 (18.8) |
| Sensory deafness and hearing impairment | 27,344 (13.9) |
| Anxiety disorders | 25,373 (12.9) |
| Stroke or Transient Ischemic Attack (TIA) | 24,287 (12.3) |
| Alzheimer’s disease and related or senile dementia | 15,361 (7.8) |
| Acute myocardial infarction | 8,117 (4.1) |
| Migraine and other chronic headache | 5,444 (2.8) |
| Mobility impairments | 3,698 (1.9) |
| Epilepsy | 2,914 (1.5) |
| Bipolar disorder | 2,637 (1.3) |
| Schizophrenia and other psychotic disorders | 2,043 (1.0) |
| Traumatic Brain Injury (TBI) & nonpsychotic mental disorders due to brain damage | 1,533 (0.8) |
| Personality disorders | 970 (0.5) |
| Spinal cord injury | 664 (0.3) |
| Attention-Deficit/Hyperactivity Disorder and other conduct disorders (ADHD) [b] | 638 (0.3) |
| Multiple sclerosis and transverse myelitis | 543 (0.3) |
| Sensory blindness and visual impairment | 306 (0.2) |
| Muscular dystrophy | 63 (0.0) |

%=Percentage

[a] The frequencies and proportions for the presence of clinical conditions are displayed in the table, and those for the absence of conditions are omitted.

[b] We kept the label for ADHD as assigned by the CCW although ADHD may no longer be considered a conduct disorder by many professional societies.

| Clinical Conditions | Difference in mean Euclidean distance | Lower 95% CL | Upper 95% CL |
|---|---|---|---|
| Cataracts | -1.1 | -1.3 | -1.0 |
| Ischemic heart disease | -0.7 | -0.8 | -0.6 |
| Rheumatoid/Osteoarthritis | -1.0 | -1.1 | -0.9 |
| Diabetes | -0.5 | -0.6 | -0.4 |
| Glaucoma | -0.8 | -0.9 | -0.7 |
| Heart failure | -0.8 | -1.0 | -0.7 |
| Acquired hypothyroidism | -0.8 | -0.9 | -0.7 |
| Peripheral vascular disease | -1.1 | -1.2 | -1.0 |
| Chronic Obstructive Pulmonary Disease (COPD) | -0.8 | -0.9 | -0.7 |
| Chronic kidney disease | -0.5 | -0.6 | -0.4 |
| Chronic pain fatigue and fibromyalgia | -0.7 | -0.9 | -0.6 |
| Depression | -0.8 | -0.9 | -0.7 |
| Sensory deafness and hearing impairment | -0.7 | -0.8 | -0.6 |
| Anxiety disorders | -0.7 | -0.9 | -0.6 |
| Stroke or Transient Ischemic Attack (TIA) | -0.9 | -1.0 | -0.7 |
| Alzheimer's disease and related or senile dementia | -1.2 | -1.4 | -1.0 |
| Acute myocardial infarction | -0.6 | -0.8 | -0.3 |
| Migraine and other chronic headache | -0.6 | -0.9 | -0.3 |
| Mobility impairments | -0.9 | -1.2 | -0.6 |
| Epilepsy | -0.9 | -1.3 | -0.6 |
| Bipolar disorder | -0.4 | -0.8 | 0.0 |
| Schizophrenia and other psychotic disorders | -0.9 | -1.4 | -0.5 |
| TBI and nonpsychotic mental disorders due to brain damage | -1.0 | -1.5 | -0.5 |
| Personality disorders | 0.2 | -0.5 | 1.0 |
| Spinal cord injury | -0.8 | -1.5 | -0.1 |
| ADHD and other conduct disorders | 0.4 | -0.4 | 1.3 |
| Multiple sclerosis and transverse myelitis | -0.4 | -1.3 | 0.4 |
| Sensory blindness and visual impairment | -2.1 | -2.8 | -1.4 |
| Muscular dystrophy | -1.7 | -2.9 | -0.5 |

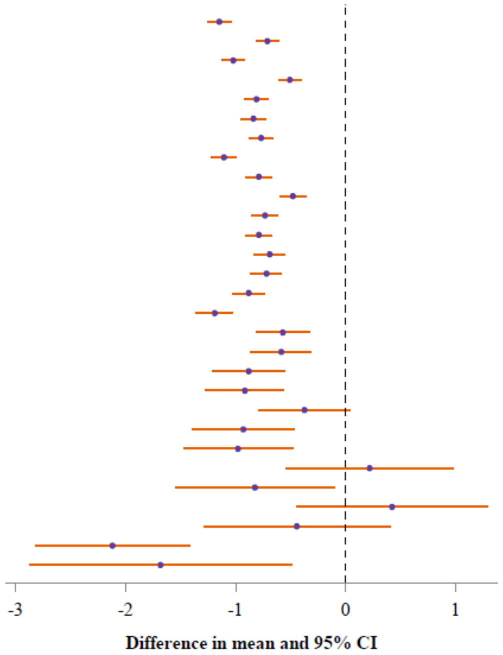


Supplementary Figure S1. Forest plot of differences in mean Euclidean distance from residential address to crash location and associated 95% confidence intervals (CI) among crash-involved drivers in New Jersey with and without the clinical conditions, ages 68 years and older, 2007-2017. Each difference in mean was found by subtracting the mean Euclidean distance of the group without the clinial condition from the group with it.

CI=Confidence Interval, CL=Confidence Limit, TBI=Traumatic Brain Injury, ADHD=Attention-Deficit/Hyperactivity Disorder
